# Assessing the Relationship Between Cannabis Use and DKA Outcomes: Insights from a National Inpatient Sample Study

**DOI:** 10.1101/2024.07.12.24310331

**Authors:** Aastha Randhawa, Kundan Jana, Moin Sattar

**Affiliations:** Department of Medicine, Maimonides Medical Center

## Abstract

DKA hospitalizations in the United States have risen across all age groups, with the highest rates observed in individuals under 45 years of age. While the ongoing decline in in-hospital DKA mortality is encouraging, further efforts are needed to identify at-risk populations. Cannabis is one of the most used recreational drugs in the world, and studies have shown that it may be related to an increased risk of DKA. While prior data assessed it impact on incidence on DKA, there exists a knowledge gap about its impact on in-patient outcomes of DKA. We designed a study aimed to investigate the impact of cannabis use on DKA outcomes using the National Inpatient Sample (NIS) dataset. Our study is a retrospective observational study, in which, we identified patient with DKA and divided them into two cohorts: those who use cannabis and those who don’t. After matching both the groups for age groups, medical conditions (e.g., AIDS, COPD, heart failure, hypertension, liver disease, malignancy), gender, obesity, smoking status, and race/ethnicity, the two groups were compared for the primary outcome of inpatient mortality and secondary outcomes including shock, acute kidney injury, acute coronary syndrome, ischemic stroke, acute respiratory failure, invasive mechanical ventilation, cardiac arrest. Contrary to prior studies showing increased overall mortality risk associated with cannabis use, out study does not show worse DKA outcomes in cannabis users. In fact, cannabis is associated with a statistically significant decrease in the odds of mortality in the matched cohort. While no conclusion regarding causative or protective effects could be drawn from this study, it helps explore unique demographic and clinical characteristics of populations, to identify at-risk populations for various outcomes of in-patient DKA admissions.

## INTRODUCTION

Diabetic ketoacidosis (DKA) is a condition which occurs due to an absolute or relative deficiency of insulin leading to uncontrolled hyperglycemia, ketonemia and metabolic acidosis.^1^DKA occurs most frequently in patients with type 1 diabetes, commonly precipitated by infections, eating disorders, or any other physiological stressor. ^2^

Unfavorable clinical outcomes include renal dysfunction, cardiac arrest, and respiratory failure to name a few, with cerebral edema being the most feared complication. Severe comorbidities such as myocardial infarction, overwhelming sepsis, or acute pancreatitis are known to be associated with worse outcomes.^1,2^

Prior data suggested that although in-hospital fatality rate declined steadily in the last two decades, the incidence of DKA continues to rise, most notably in persons aged <45 years, which is also the age group associated with high cannabis use. DKA is also has a high financial burden on healthcare with the direct and indirect annual cost of DKA hospitalization being around $5.1 billion. ^3,4^

Cannabis (marijuana) refers to the dried leaves, flowers, stems, and seeds from the *Cannabis sativa* or *Cannabis indica* plant. It is one of the most commonly used psychotropic drug in the world, with an estimated 200 million users worldwide.^5^ Despite this, there is limited research on the influence of cannabis use on DKA outcomes. This study aims to investigate the impact of cannabis use on DKA outcomes using the National Inpatient Sample (NIS) dataset.

## METHODS

This is a retrospective observational study using the National Inpatient Sample (NIS) database. This dataset represents the largest publicly available all-payer inpatient healthcare database designed to produce U.S. regional and national estimates of inpatient utilization, access, cost, quality, and outcomes. This study is IRB exempt, given the NIS uses anonymized public-use data without personally identifiable information.

We identified patients with DKA using the following ICD codes: “E1010”,“E1011”,“E1110”,“E1111”. The identified patients were divided into two cohorts: those who use cannabis and those who don’t. Comorbidities such as hypertension, obesity, chronic respiratory failure, heart failure were also accounted for.

Finally, the two groups were compared for the primary outcome of inpatient mortality and secondary outcomes including shock, acute kidney injury, acute coronary syndrome, ischemic stroke, acute respiratory failure, invasive mechanical ventilation, cardiac arrest.

Baseline categorical data were compared using Rao-Scott chi square test, and continuous data were compared using Student’s *t*-test. Identification of risk factors was done using logistic regression models with inverse probability weighting for complex survey data. Covariates were assessed using univariate regression analysis, and factors with *p* value less than 0.1 in the univariate analysis were entered in the final model. The univariate and multivariable associations of presumed risk factors of mortality in DKA with Cannabis Use Disorder patients were performed using multivariate regression analysis.

Propensity score matching was done using the MatchIt package in R (version 4.3.0). 1:1 nearest neighbor matching was done with propensity scores estimated through logistic regression, in which occurrence of Cannabis use disorder was regressed according to other patient characteristics. Subsequently, outcomes like in-patient mortality, cerebral edema, shock, arrhythmia, acute coronary syndrome, stroke, acute renal failure, invasive mechanical ventilation, Cardiac arrest, length of stay and cost utilization were studied in the matched model using logistic regression. Data extraction with ICD-10 codes was done using Python (version 3.9.1). Statistical analysis was carried out using R (version 3.6.2 R Foundation for Statistical Computing, Vienna, Austria).

**Table 1.** Comparison of baseline characteristics between patients with and without Cannabis Use Disorder.

|  | **Cannabis Use Disorder** | **No Cannabis Use Disorder** |
| --- | --- | --- |
| Total | 20237 | 301691 |
| Median Age | 32 | 45 |
| Mean age | 34.6±12.3 | 45.5±18 |
| Male | 151592 | 12434 |
| Female | 7801 | 150057 |
| Median LOS | 3 | 3 |
| Mean LOS | 3.54±4.46 | 5.08±7.34 |
| Charlson Comorbidity Index (CCI) | 2.73±1.84 | 3.68±2.76 |
| Race: |  |  |
| Caucasian | 10550 (52.13%) | 167645 (55.57%) |
| African-American | 5836 (28.83%) | 71809 (23.80%) |
| Hispanic | 2412(11.91%) | 38954(12.9%) |
| Asian | 132 (0.65%) | 4753 (1.58%) |
| Native American | 279 (1.38%) | 3087 (1.02%) |
| Others | 514 (2.54%) | 7841 (2.60%) |
| Median cost | 6692 | 7890 |
| Pay: |  |  |
| Medicare | 2846 | 84665 |
| Medicaid | 9911 | 93834 |
| Private insurance | 4009 | 79964 |
| Self pay | 2692 | 31606 |
| No charge | 177 | 2120 |
| Others | 574 | 8991 |
| Comorbidities: |  |  |
| Chronic resp failure | 2551 | 39423 |
| COPD | 838 | 19862 |
| CHF | 879 | 31404 |
| CKD | 1835 | 42457 |
| ESRD | 644 | 16991 |
| HTN | 8669 | 158401 |
| Smoker | 9725 | 68054 |
| Obesity | 1558 | 45208 |
| HLD | 4266 | 97892 |

## RESULTS

Out of the total numbers of patients identified with DKA, 20237 patients were cannabis users and were assigned to the study group and the remaining 301691 patients did not have cannabis use disorder. Median age was found to be lower (32 years) in the group that has cannabis use disorder, as opposed to those who don’t (45 years).In the study group, the age distribution was as follows: 36.83% were under 30, 25.61% were 30-39, 24.28% were 40-49, 9.66% were 50-59, 3.99% were 60-69, 0.46% were 70-79, and 0.17% were 80 or older. In the control group, 2.89% were under 30, 1.82% were 30-39, 1.05% were 40-49, 0.65% were 50-59, 0.27% were 60-69, 0.03% were 70-79, and less than 0.01% were 80 or older. The study group had a significantly higher percentage of younger patients, while the control group had a higher percentage of older patients. Control group had a predominance of males with 61.44% males and 38.55% females, while the study group had almost equal percentages of males(50.25%) and females (49.73%). Mortality rates of unmatched data were higher in non-cannabis users (3.66%) as opposed to the study group (0.57%). Median length or stay was the same in both groups. The study group has a higher percentage of African-American (28.83%) and Native American (1.38%) participants compared to the control group (23.80% and 1.02%, respectively). Conversely, the control group has a higher percentage of Caucasian (55.57%) and Asian (1.58%) participants compared to the study group (52.13% and 0.65%, respectively). The percentages of Hispanic and “Other” participants are fairly similar between the two groups.

In the study group of 20,237 patients, the payment methods were distributed as follows: 14.06% had Medicare, 48.98% had Medicaid, 19.80% had private insurance, 13.30% were self-pay, 0.87% had no charge, and 2.84% had other modes of payment. In the control group of 301,691 patients, the distribution was: 28.06% had Medicare, 31.09% had Medicaid, 26.51% had private insurance, 10.48% were self-pay, 0.70% had no charge, and 2.98% had other modes of payment. The study group had a significantly higher percentage of Medicaid patients compared to the control group, while the control group had a higher percentage of Medicare and private insurance patients.

Charlson Comorbidity Index (CCI), which estimates the risk of death from comorbid disease was higher in the control group. ^6^ In the study group 12.60% had chronic respiratory failure, 4.14% had COPD, and 4.34% had CHF. In the control group 13.07% had chronic respiratory failure, 6.58% had COPD, and 10.41% had CHF. The study group had a similar percentage of chronic respiratory failure compared to the control group, but a lower percentage of patients with COPD and CHF. In the study group of 20,237 patients, 9.07% had CKD and 3.18% had ESRD. In the control group, about 14.07% had CKD and 5.63% had ESRD. The study group had a lower percentage of patients with both CKD and ESRD compared to the control group. In the study group, 42.84% had hypertension (HTN). The study group had a lower percentage of patients with hypertension compared to the control group. In the study group were smokers, 7.70% were obese, and 21.08% had hyperlipidemia (HLD). In the control group, were smokers, 14.98% were obese, and 32.46% had hyperlipidemia. The study group had a significantly higher percentage of smokers (48.05%) compared to the control group(22.55%). However, the percentage of obese patients was lower in the study group compared to the control group. Similarly, the percentage of patients with hyperlipidemia was lower in the study group compared to the control group. This suggests that smoking was more prevalent among the study group, while obesity and hyperlipidemia were more prevalent among the control group.

**Table 2:** Matched data for primary and secondary outcomes.

|  | **Cannabis Use** | **No cannabis Use** |
| --- | --- | --- |
| Total | 19715 | 19715 |
| Died | 112 | 216 |
| Invasive mechanical ventilation | 624 | 842 |
| Shock | 483 | 770 |
| Arrest | 116 | 154 |
| Arrhythmia | 697 | 752 |
| ACS | 490 | 489 |
| AKI | 7716 | 8147 |
| Stroke | 16 | 12 |
| Cerebral edema | 29 | 42 |
| Median LOS | 3 | 3 |

After matching both the groups for age groups, medical conditions (e.g., AIDS, COPD, heart failure, hypertension, liver disease, malignancy), gender, obesity, smoking status, and race/ethnicity, both groups had a total of 19,715 patients each.The primary outcome was to assess the mortality rate, which was found to be lower among cannabis users (0.57%) compared to non-users (1.10%).

Patients with Cannabis Use Disorder appeared to have better outcomes in terms of invasive mechanical ventilation(3.17% vs. 4.27%), shock(2.45% vs. 3.91%), cardiac arrest (0.78% vs. 0.59%), acute kidney injury (39.22% vs. 41.31%), or similar outcomes in terms of arrhythmia, and cerebral edema compared to those without CUD.

The median length of stay and the median cost was the same for both groups. Only the incidence of stroke was slightly higher in the CUD group, but the difference was minimal. These results suggest that CUD might not negatively impact these particular health outcomes.

**Table 3:** Multivariate analysis-Matched Data for Mortality.

|  | Odds Ratio | Low CI | Upper CI | p-value |
| --- | --- | --- | --- | --- |
| CANNABIS | 0.9972 | 0.9957 | 0.9987 | <0.01 |
| ACS | 1.0055 | 0.9929 | 1.0183 | 0.39 |
| ARREST | 1.6274 | 1.5326 | 1.7281 | <0.01 |
| ARRHYTH | 1.0094 | 0.9998 | 1.019 | 0.06 |
| SHOCK | 1.1079 | 1.0882 | 1.128 | <0.01 |

Multivariate analysis was used to assess the impact of various factors on mortality in the matched cohort. The odds ratio for Cannabis Use indicates a slightly reduced risk of mortality (OR < 1). This means that having CUD is associated with a statistically significant decrease in the odds of mortality in the matched cohort. Other factors like cardiac arrest and shock significantly increase the risk of mortality.

## DISCUSSION

DKA is known to have the highest rate in younger adults <45 years of age. This is also the age group known for higher rates of cannabis use. ^7,8^ There have been prior studies that have shown an association between cannabis use and a increased risk of DKA in adults with type 1 diabetes.^9,10^ Cannabis hyperemesis syndrome (CHS) has also been linked to repeated episodes of DKA in individuals with type 1 diabetes. ^11^ A recent study reported that marijuana use was associated with lower levels of fasting insulin and insulin resistance. ^12^ However, to the best of our knowledge, no study has been done to compare in-patient hospital outcomes of DKA in cannabis users vs non users.

We discovered that the study group of DKA patients who used cannabis comprised a higher percentage of males compared to the control group. Numerous studies have demonstrated that males are nearly twice as likely to use cannabis as females. ^13,14^Among major ethnic groups, Black Americans have the highest rate of cannabis use, followed by Whites and Hispanics. ^15^ A similar distribution of ethnic groups has also been noted in our study. Our unmatched study group exhibited other differences in baseline characteristics, such as a higher percentage of smokers. This may be attributed to the prevalence of concurrent high-risk behaviors in individuals. ^16^

Contrary to expectations, obesity was significantly lower in the study group. While previous studies have shown mixed evidence on this topic, the majority of recent literature indicates that cannabis use is associated with reduced obesity and lower waist circumference. Additionally, state-level legalization of medical cannabis has been linked to a rapid decline in obesity rates.^12,17,18^

In our study, the Charlson Comorbidity Index (CCI) was higher in the control group of the unmatched data. So, both the groups were matched based on various criteria, including age groups, medical conditions (e.g., AIDS, COPD, heart failure, hypertension, liver disease, malignancy), gender, obesity, smoking status, and race/ethnicity, before comparing outcomes. Primary outcome of this study was to assess the inpatient mortality rate between both groups. The relationship between cannabis use and specific causes of mortality in the general population is still largely unexamined. Many prior studies indicate an increased risk of all-cause mortality associated with cannabis use.^19,20^There have been studies showing increased risk of cardiovascular events related to cannabis use started in younger age. ^21^In another large study, the relative risk of death among high cannabis users was significantly higher compared to non-users, but no excess mortality was found after adjusting for social background variables in a multivariate model. ^22^ A large cohort study using UK Biobank demonstrated a positive association between CVD mortality and heavy lifetime cannabis use, but only among females. ^23^In our study, mortality rates were 0.57% for cannabis users and 1.10% for non-users for unmatched data. After using Propensity matching, mortality rate was still lower in cannabis users. Both groups were matched for age ruling out the possibility of age difference playing a role. Both propensity scoring and multivariate analysis were used to reduce the impact of confounders such as comorbidities, thereby providing a clearer understanding of the true relationship between the cannabis use and the outcome of DKA.

Some studies have identified temporal associations between marijuana use and serious adverse events, including myocardial infarction, sudden cardiac death, cardiomyopathy, stroke, transient ischemic attack, and cannabis arteritis.^24–26^ A recent study suggests that cannabis use may independently increase the risk of myocardial infarction and stroke, with more frequent use linked to a higher likelihood of adverse cardiovascular outcomes, regardless of the method of consumption.^27^ In our study, we found no such difference between the two groups.

DKA is known to cause electrolyte imbalances such as hypokalemia and hypomagnesemia. The highest risk of hypokalemia occurs within the first 48 hours of admission, and severe hypokalemia is associated with increased mortality.^4^ These electrolyte imbalances can lead to cardiac arrhythmias. In comparing the two groups for this outcome, we did not find a significant difference in the incidence of cardiac arrest or arrhythmias. Another outcome studied was acute kidney injury (AKI), a recognized complication of DKA that independently increases mortality risk among hospitalized DKA patients. ^28^ Our study depicted similar outcomes in terms of renal failure in both the groups.

Despite establishing better outcomes of DKA in cannabis users compared to non-users, since this study is based on a population dataset, it inherently lacks the ability to establish causal or protective effects due to several limitations. Firstly, it cannot account for the severity of conditions or factors that are not coded in the database, leading to potential misclassification or incomplete data. Secondly, retrospective studies are observational and thus susceptible to various biases, such as selection bias and recall bias. These studies can identify associations but cannot definitively prove causation due to the absence of randomization. Additionally, the study design cannot control for all potential confounding variables that might influence the outcomes. As a result, while population-based retrospective studies can provide valuable insights and generate hypotheses, they are not robust enough to establish causative or protective effects conclusively.

In conclusion, the hospitalization rate for DKA in individuals with diabetes under the age of 45 was much higher than in those aged 65 and older. Consequently, to address the rising number of DKA hospitalizations, it is crucial to consider the unique demographic and clinical characteristics of younger populations, including youths and young adults. Despite the encouraging ongoing decline in in-hospital DKA mortality, further efforts are needed to identify at-risk populations. Understanding these factors is essential for developing targeted interventions and preventative measures.

## Data Availability

This is a retrospective observational study using the National Inpatient Sample (NIS) database. This dataset represents the largest publicly available all-payer inpatient healthcare database designed to produce U.S. regional and national estimates of inpatient utilization, access, cost, quality, and outcomes.

https://hcup-us.ahrq.gov/nisoverview.jsp

---

**Fig 1:**
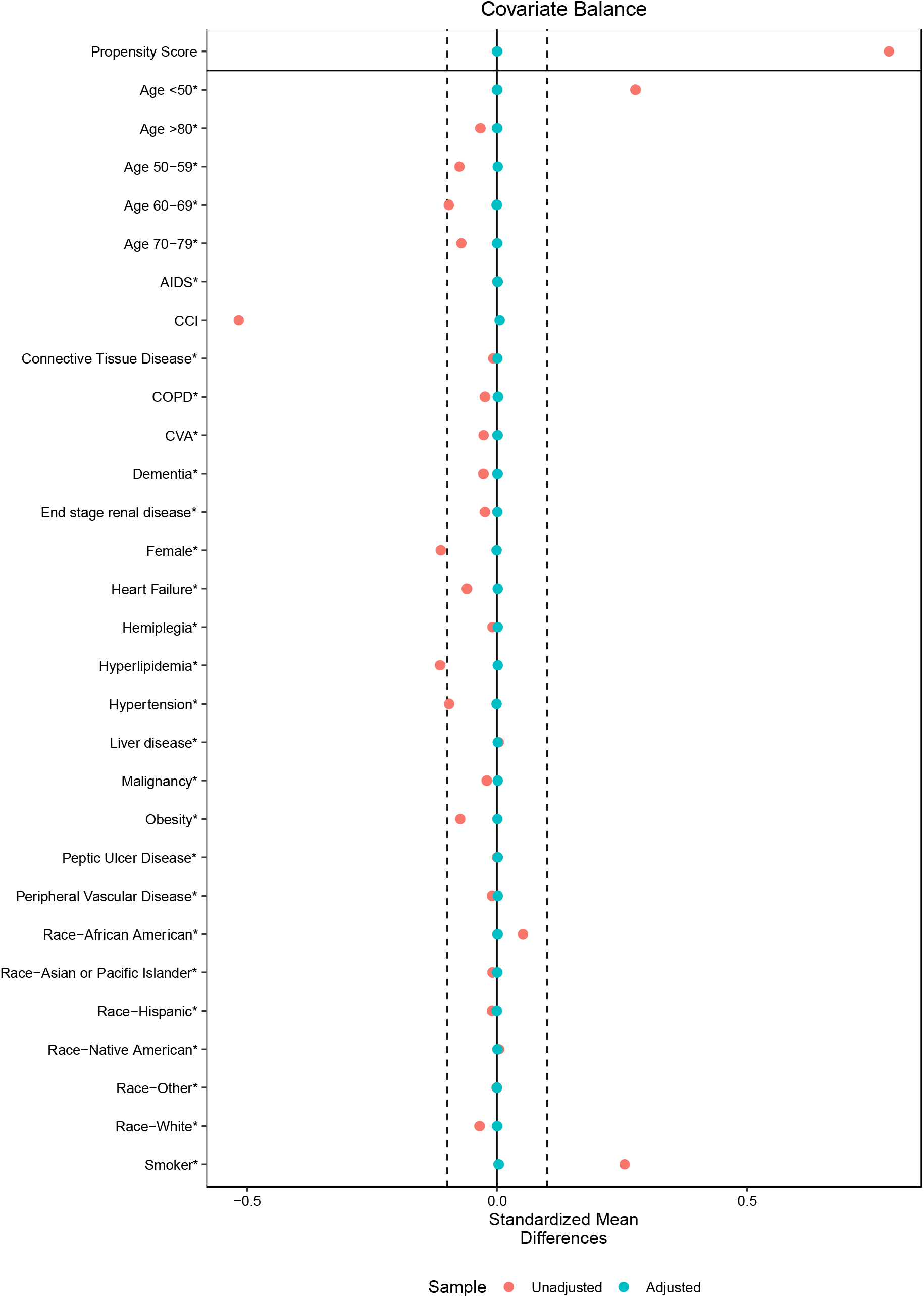
Propensity score matching

